# Factors associated with late-stage cervical cancer diagnosis in Northwest Ethiopia: A cross-sectional study

**DOI:** 10.64898/2026.09.25.26364043

**Authors:** Eyaya Misgan Asress, Muluken Azage Yenesew, Dabere Nigatu, Awoke Derbie, Daniel A. Enquobahrie, Tesfaye Mersha, Tsion Zewdu Minas, Robel Alemu, Tsegaselassie Workalemah

**Affiliations:** Department of Obstetrics and Gynaecology, College of Medicine and Health Sciences, Bahir Dar University, Bahir Dar, Ethiopia; Department of Obstetrics and Gynaecology, College of Medicine and Health Sciences, University of Rwanda, Kigali, Rwanda; Department of Environmental Health, School of Public Health, College of Medicine and Health Sciences, Bahir Dar University, Bahir Dar, Ethiopia; Department of Reproductive Health and Population Studies, School of Public Health, College of Medicine and Health Sciences, Bahir Dar University, Bahir Dar, Ethiopia; Department of Medical Microbiology and Immunology, College of Medicine and Health Sciences, Bahir Dar University, Bahir Dar, Ethiopia; Department of Epidemiology, School of Public Health, University of Washington, Seattle, WA; Indiana University School of Medicine, Indianapolis, Indiana, USA; Department of Pathology, School of Medicine, Johns Hopkins University, Baltimore, MD; Department of Obstetrics and Gynaecology, College of Medicine and Health Sciences, University of Gondar, Gondar, Ethiopia; University of Utah, Salt Lake City, Utah, USA

**Keywords:** Cervical cancer, FIGO stage, Risk factor, Predictors, Hospital, Cervical cancer symptoms, Ethiopia

## Abstract

**Background:** Cervical cancer (CC), mostly associated with high-risk human papillomaviruses (hrHPV) infection, is the most common cancer that disproportionately affects women in developing countries, including Ethiopia. Early detection of CC and initiation of treatment are key to positive health outcomes. Identifying factors associated with delayed CC diagnosis is essential for effective prevention and control efforts. This study assessed the stage at diagnosis and examined associated clinical and sociodemographic factors.

**Methods:** An institution-based cross-sectional study was conducted among 284 women newly diagnosed with CC who attended Tibebe Ghion Specialized Hospital (TGSH) between 1^st^ January 2021 and 31^st^ December 2022. A structured questionnaire was used to collect data on the participants’ demographic, reproductive, and gynaecologic history, and clinical profiles. CC stage at diagnosis was categorised as early (stage I-IIA) or late (stage IIB-IV). Logistic regression models (unadjusted and adjusted) were used to examine associations of sociodemographic or clinical factors with stage at CC diagnosis.

**Results:** The mean age (±standard deviation) at presentation was 53.2 ± 11.3 years, with an age range of 18 – 80 years. Most of the patients (73.6%) were from rural areas, and three-fifth (58.1%) had no medical insurance. Among participants, 74.6% (95% CI: 69.4, 79.6%) had late-stage CC. The odds of late-stage CC diagnosis were higher for women from rural areas (AOR: 2.21; 95% CI:1.18, 4.12; p<0.013), and those with anaemia (defined as <12 mg/dl haemoglobin) (AOR: 3.42; 95% CI: 1.88, 6.23; p<0.001). On the other hand, the odds of late-stage CC diagnosis were lower in HIV positive women (AOR:0.41; 95%CI:0.19, 0.88; p<0.021).

**Conclusion:** The proportion of late-stage diagnosis among CC patients was high. Residential area, anaemia status, and HIV serostatus were factors associated with stage at CC diagnosis. These findings can inform screening efforts of CC and call for expanding of the screening services to the rural setting in order to reduce the burden of late-stage CC diagnosis.

## Introduction

Cervical cancer (CC) is a disease in which the cells of the cervix start to grow uncontrollably. This abnormal growth could arise from different predisposing factors such as having multiple sexual partners, early onset of sexual intercourse, history of cigarette smoking, having a male sexual partner with a history of multiple sexual partners, family history of cancer, and, importantly, human papillomavirus (HPV) infections. Persistent infection with high-risk HPV, typically over years to decades, can lead to progressive cervical epithelial changes from normal cells to cervical intraepithelial neoplasia (dysplasia). If untreated, high-grade lesions may progress to carcinoma in situ and eventually to invasive cervical cancer [1, 2].

CC is the fourth most frequently diagnosed cancer and the fourth leading cause of cancer mortality among women, with 604,000 new cases and 342,000 deaths worldwide in 2020 [1]. It is also the second most prevalent cancer in women aged between 15 and 44 years worldwide. [1, 3, 4]. As a major public health problem, CC affects the most vulnerable groups, such as women in rural areas and of poor socioeconomic status, and women living with human immunodeficiency virus (HIV) [5, 6]. The incidence and mortality from CC are the highest (10-fold higher) in low- and middle-income countries, particularly in sub-Saharan Africa. [7]. Sub-Saharan countries, including Ethiopia, fall in this category [1, 3, 4].

In Ethiopia, CC is the second most diagnosed cancer and the leading cause of cancer death in women, with about 7,445 newly diagnosed cases and 5,338 deaths every year. Most CC patients in Ethiopia seek healthcare at an advanced stage, when the effectiveness of treatment is limited. [8]. As a major public health problem in Ethiopia, CC disproportionally affects the country’s vulnerable populations, particularly individuals with limited access to health care, low levels of education, residents of rural communities with poor awareness of screening services, and those with biological risk factors such as HIV infection [9].

One of the most important prognostic factors is stage at diagnosis, which is strongly correlated with better chances of survival. However, there is a dearth of evidence on the magnitude and predictors of late-stage presentation of CC patients in Ethiopia. Therefore, the study aimed to describe sociodemographic, socioeconomic, clinical, and histopathologic profiles of CC patients, and assess the stage at CC diagnosis (and associated factors) among patients who attended Tibebe Ghion Specialized Hospital, Ethiopia. Findings will inform guidelines to support healthcare professionals involved in CC prevention and treatment. The findings may also be used to guide future research and policymaking.

## Methods and materials

### Study design and setting

This is an institution-based cross-sectional study conducted at Tibebe Ghion Specialized Hospital (TGSH). TGSH is a government-owned referral teaching hospital under the Federal Ministry of Health, located in Bahir Dar city, Ethiopia. Bahir Dar is the capital city of the Amhara region, located 565 Kilometres from Addis Ababa, the capital city of Ethiopia. The hospital is a teaching hospital that has been operating since 2018, providing diagnostic and treatment services for more than ten million people.

### Study population

The study was conducted among inpatient and outpatient CC patients who were evaluated and treated in the gynaecologic-oncology and clinical oncology unit from January 1, 2021, to December 31, 2022. Cervical cancer patients with a pathological diagnosis of CC and aged 18 years and older were eligible for the study. All CC patients who met the predefined eligibility criteria were included until the required sample size (based on pre-study power calculations) was achieved. Patients with incomplete medical records were excluded from the study (N=96). The final analytic population included N=284. The Institutional Review Board of the College of Medicine and Health Sciences (IRB-CMHS), Bahir Dar University (BDU), approved the study under protocol number 655/2023. We also obtained a letter of permission from TGSH. The study relies on chart review. In the data collection checklists, the names of the patients and their medical record numbers were not included for the sake of their privacy. Hence, the authors had no access to information that could identify individual participants during or after data collection. All the processes of the research were performed and secured in accordance with the relevant guidelines and regulations.

### Data collection tools and methods

A structured questionnaire was prepared to collect the socio-demographic, socio-economic, and clinical characteristics of patients (S1 File) [10–16]. Data were abstracted by reviewing the CC patients’ medical records. For data abstraction, the CC patients’ charts were accessed from February 20, 2023 to June 20, 2023. The data collectors were two trained midwives. The data collection was supervised by two fourth-year obstetrics and gynaecology residents.

This manuscript is written according to the “Strengthening the Reporting of Observational Studies in Epidemiology (STROBE)” guideline (S1 Checklist).

### Study variables and measurement

The dependent variable is the stage at diagnosis of cervical cancer. Stage at diagnosis was classified according to the FIGO CC stage classification: The early stage is defined as patients presenting with FIGO stages I-IIA. In contrast, the late stage at presentation is defined as patients presenting with FIGO stages IIB-IV[11]. The presenting complaint was the symptom that was identified at the time of first evaluation (the ‘first attributed’ symptom), the patient reported and documented in the chart as the chief complaint [17]. Patients’ haemoglobin level below 12.0 g/dl was classified as anaemic [18]. Co-morbidity was characterized by the presence of any conditions mentioned in the Carlson comorbidity Index [19] other than CC at diagnosis.

### Data analysis

Data were entered into EP-info version 6.4.1, cleaned and coded, and then transferred to SPSS version 25 for analysis. Descriptive summaries, including frequencies, proportions, percentages, means, and standard deviations, were computed and presented in tables. Binary logistic regression analysis was used identify predictors of late-stage cervical cancer at diagnosis. Both unadjusted and adjusted models were fitted. In the unadjusted model, each independent variable was analysed individually with the outcome variable. Variables with *p* < 0.05 in the unadjusted analysis were selected to compute the adjusted logistic regression model to identify factors associated with late-stage CC diagnosis. Odds Ratios (ORs) with 95 % confidence interval were calculated. Significant association was declared at p-value less than 5%.

## Results

### Demographic and reproductive characteristics of CC patients

A total of 284 CC patients were included in the study. The mean age (±standard deviation) at presentation was 53.2 ± 11.3 years, with an age range of 18 – 80 years. Most of the patients (N=209,73.6%) were from rural areas, and more than half (N=152, 53.7%) were married. three-fifth of patients (N=165, 58.1%) had no medical insurance. Three-fourths of the patients (N=211, 74.3%) were post-menopausal. Furthermore, 23.2%, 15.5%, and 71.8% were hormonal contraceptive users, HIV positive, and had a history of treatment for sexually transmitted infections, respectively. One hundred and seventy-eight (62.7%) patients had delivered five or more times in their lifetimes, with a mean number of children of 6.2 (Table 1).

**Table 1:** Selected demographic and reproductive characteristics of CC patients, TGSH, Bahir Dar, Northwest Ethiopia, 2023.

| Characteristics | Response options | Number (%) |
| --- | --- | --- |
| Age groups (in years) | Mean, SD | 53.2± 11.3 |
|  | 30-40 | 47 (16.5) |
|  | 41-50 | 90 (31.7) |
|  | 51-60 | 85 (29.9) |
|  | >60 | 62 (21.8) |
| Permanent residence | Urban | 75 (26.4) |
|  | Rural | 209 (73.6) |
| Marital status | Married | 152 (53.5) |
|  | Divorced | 47 (16.6) |
|  | Widowed | 45 (15.8) |
|  | Unknown | 40 (14.1) |
| Number of deliveries | Mean, SD | 6.2±2.6 |
|  | <5 | 106 (37.3) |
|  | ≥5 | 178 (62.7) |
| Use of a hormonal contraceptive | Yes | 66 (23.2) |
|  | No | 218 (76.8) |
| Menopausal status | Premenopausal | 73 (25.7) |
|  | Postmenopausal | 211 (74.3) |
| Health insurance | Insured CBHI | 119 (41.9) |
|  | Not insured | 165 (58.1) |
| HIV serostatus | Positive | 44 (15.5) |
|  | Negative | 240 (84.5) |
| History of treatment for vaginal discharge | Yes | 204 (71.8) |
|  | No | 80 (28.2) |
| History of cervical screening | Yes | 79 (27.8) |
|  | No | 205 (82.2) |

SD Standard deviation, CBHI Community-based health insurance

### Clinical characteristics of CC patients

All patients (100%) presented with symptoms related to CC, with the most common symptoms being vaginal bleeding (45.4%) and vaginal discharge (36.6%) (Table 2). The median duration of presenting symptoms was 6 months (range: 0.5-84 months). The patients had visited at least two health facilities before reaching TGSH, and the majority (N=255, 89.8%) visited three or more health care facilities. About nine in ten (N=245, 86.3%) patients were referred to TGSH with a suspected diagnosis of CC.

**Table 2:** Clinical characteristics of CC patients, TGSH, Bahir Dar, Northwest Ethiopia, 2023.

| Characteristics | Response options | Number (%) |
| --- | --- | --- |
| Presenting complaint | Vaginal discharge | 104(36.6) |
|  | Vaginal bleeding | 129(45.4) |
|  | Pain (abdominal or pelvic pain) | 51(18) |
| Duration of chief complaint in months | < 3 months | 92(32.2) |
|  | 3 months to 6 months | 60(21.2) |
|  | > 6 months | 132(46.5) |
| Number of health facilities visited | Two | 29(10.1) |
|  | Three or more | 255(89.8) |
| Referral diagnosis | Cancer suspected/confirmed | 245(86.3) |
|  | Cancer not suspected | 39(13.7) |
| Size of cervical mass | Mean, SD | 4.2±1.7 |
|  | <4 cm | 97(34.2) |
|  | ≥4 cm | 187(65.8) |
| Vaginal involvement | Not involved | 148(52.1) |
|  | Involved | 136(47.9) |
| Parametrium | Not involved | 179(63) |
|  | Involved | 105(37) |
| Hydronephrosis | No | 233(82.1) |
|  | Yes | 51(17.9) |
| Histological type | Squamous cell carcinoma | 261(91.9) |
|  | Adenocarcinoma | 23(8.1) |
| FIGO stage | Stage I | 62(21.8) |
|  | Stage II | 76(26.8%) |
|  | Stage III | 91(32%) |
|  | Stage IV | 55(19.4%) |
| Hemoglobin in mg/dl | Mean ±SD | 12.4±2.2 |
|  | <12 gm/dl | 100(35.2) |
| | ≥12 gm/dl | 184(64.8) |
| Comorbidity | Any comorbidity | 82(28.9) |
|  | No comorbidity | 202(71.1) |
| Type of initial CC treatment | Surgery | 69(24.5) |
|  | Chemo radiation | 215(75.5) |

On clinical pelvic examination, a palpable cervical mass was detected in all but one patient (N=283, 99.6%). The mean diameter of the cervical mass was 4.2±1.7 cm (range: 0 – 9 cm). Two-thirds of the patients (N=187, 65.4%) had cervical mass size ≥4 cm. In addition, involvement of the vagina was reported among 136 (47.9%) patients, while the parametrium was involved in 105 (37%), and ultrasound documented hydronephrosis was reported in 51 (17.9%) patients. Most patients (N=261, 91.9%) had cervical punch biopsy histopathology report of squamous cell carcinoma. Three-quarters (N=212, 74.6%, 95% CI: 69.4, 79.6%) of CC patients were diagnosed at late stages (IIB-IV). One in five (N=62, 21.8%) CC patients were in stage I, and a quarter of the CC patients (N=76, 26.8%) were in stage II. On the other hand, one third (N=91, 32%) were diagnosed with stage III and 19.4% (N=55) at stage IV (Table 2)

### Factors associated with CC stage at diagnosis

Participants’ place of residence, parity, presenting complaint, history of use of Oral Contraceptive Pills (OCP), HIV serostatus, comorbidity, haemoglobin level in mg/dL, and cervical mass size were factors associated with stage at diagnosis of CC in univariate analyses (all p-values <0.05) (Table 3). However, in multivariable logistic regression analysis, only place of residence, HIV serostatus, and haemoglobin(mg/dL) were statistically associated with stage at CC diagnosis. The odds of late-stage diagnosis were higher for women from rural areas (AOR: 2.21; 95% CI: 1.18, 4.12; p<0.013), those with anaemia (AOR: 3.42; 95%CI:1.88, 6.23; p<0.001), and cervical mass size of ≥4 cm (AOR:5.24; 95%CI:2.93, 9.38; p<0.001). On the other hand, the odds of late stage at diagnosis were lower among HIV positive women (AOR:0.41; 95%CI:0.19, 0.88; p<0.021) (Table 4).

**Table 3:** Univariate analysis of associated factors with CC stage at diagnosis in TGSH, Bahir Dar, Northwest Ethiopia, 2023.

| Exposure variables |  | CC stage |  | COR (95%CI) |
| --- | --- | --- | --- | --- |
|  |  | Late, n (%) | Early, n (%) |  |
| Age groups | 30-40 | 31(14.6) | 16(22.2) | Reference |
|  | 41-50 | 70(33) | 20(27.8) | 0.87(0.43,1.77) |
|  | 51-60 | 62(29.2) | 23(31.9) | 1.49(0.73,3.05) |
|  | >60 | 49(23.1) | 13(18.1) | 1.54(0.73,3.32) |
| Permanent residence | Urban | 49(23.1) | 26(36.1) | Reference |
|  | Rural | 163(76.9) | 46(63.9) | 2.59(1.50,4.48) * |
| Number of children | <=5 | 70(33) | 36(50%) | Reference |
|  | >5 | 142(67) | 36(50%) | 1.66(1.02,2.69) * |
| Presenting complaint | Vaginal discharge | 69(32.5) | 35(48.6) | Reference |
|  | Vaginal bleeding | 101(47.6) | 28(38.9) | 1.83(1.02,3.28) * |
|  | Abdominal Pain | 42(19.8) | 9(12.5) | 2.37(1.04,5.41) * |
| History of use of OCP | Yes | 44(20.8) | 22(30.6) | 0.45(0.25,0.78) * |
|  | No | 168(79.2) | 50(69.4) | Reference |
| HIV serostatus | Positive | 26(12.3) | 18(25) | 0.35(0.18,0.69) * |
|  | Negative | 186(87.7) | 54(75) | Reference |
| Hemoglobin in mg/dl | <12 | 87(41) | 13(18.1) | 3.16(1.63,3.11) * |
|  | >=12 | 125(59) | 59(81.9) | Reference |
| Comorbidity | No | 158(74.5) | 44(61.1) | Reference |
|  | At least one | 54(25.5) | 28(38.9) | 1.94(1.16,3.27) * |

\* P-value<0.05 and variables associated with stage at CC diagnosis on univariate analysis
and selected for multivariable analysis, COR: crude odds ratio, CI: Confidence interval

**Table 4:** Multivariate analysis of factors associated with CC stage at diagnosis in TGSH, Bahir Dar, Northwest Ethiopia, 2023.

| Exposure variable |  | CC stage |  | COR | AOR (95%CI), p-value |
| --- | --- | --- | --- | --- | --- |
|  |  | Late, n (%) | Early, n (%) |  |  |
| Permanent residence | Urban | 49(23.1) | 26(36.1) |  | Reference |
|  | Rural | 163(76.9) | 46(63.9) | 2.59 | 2.21(1.18,4.12) |
| HIV serostatus | Positive | 26(12.3) | 18(25) | 0.35 | 0.41(0.19,0.88) |
|  | Negative | 186(87.7) | 54(75) |  | Reference |
| Hemoglobin in mg/dl | <12 | 87(41) | 13(18.1) |  | Reference |
|  | >=12 | 125(59) | 59(81.9) | 3.16 | 3.42(1.88,6.23), <0.001 |

Adjusted for age, comorbidity, parity, OCP use, and presenting complaints, CC: cervical
cancer, COR: crude odds ratio, AOR: Adjusted odds ratio, CI: Confidence interval

## Discussion

The main aim of this study was to assess the stage at diagnosis of cervical cancer and identify the associated factors in a tertiary care level hospital among symptomatic women in northwestern Ethiopia. We found out that a considerable proportion of women with cervical cancer are diagnosed at a late stage, while factors like rural residence, being anaemic, and HIV positive serostatus were found to be associated with the odds of having late-stage disease.

In the current study, the proportion of patients diagnosed with late-stage cervical cancer was 74.6% (95% CI: 69.4,79.6%). The proportion of late-stage CC in our study is lower than that reported in a cross-sectional study conducted in Addis Ababa, where 87.9% of patients were diagnosed at a late stage [10]. It is also lower than reports from studies in Zaria, Northern Nigeria (98%), Muhimbili National Hospital, Tanzania (90%), and Nepal (80.9%) [20–22]. On the other hand, our current finding is higher than the report from a multicenter cross-sectional study among residents of Addis Ababa, which reported a 60.4% proportion of late-stage diagnosis [13]. A slightly lower proportion of late-stage CC was also reported from East African countries, including North-western Tanzania (63.9%) [23], Northern Uganda (66%) [24], and Kenya (69%) [25]. A much lower proportion of late-stage CC diagnoses has been reported from Morocco (39.9%)[26], India (45.4%)[27], England (28%) [28], and Mexico (17.8%)[29]. Differences in health care delivery packages, socioeconomic disparities, study period, and study population eligibility could account for variations in the magnitude of late-stage disease across studies.

Factors such as differences in participants’ awareness of CC screening practices, availability and access to CC screening and treatment services, women’s health policies of nations, and socio-demographic factors contribute to a later stage of CC diagnosis. The majority (76.9%) of our study participants live in rural settings, and they may lack awareness and knowledge about CC, including the ways of transmission, prevention, and the symptoms for which they should seek medical attention. In a systematic review and meta-analysis by Derbie et al., the overall pooled prevalence of good knowledge about CC prevention was 43%, suggesting that Ethiopian women had poor knowledge about CC prevention[30]. The proportion of CC screening was quite low (27.8%) in our study. A meta-analysis by Desta et al. revealed that CC screening among eligible women in Ethiopia was below 15%. The lowest national screening coverage was documented in Amhara regional state, where our study area is located.[31]. All these factors could explain the greater proportion of late-stage CC in the present study. Overall, the high proportion of late-stage CC diagnoses observed in this study underscores the need to enhance community awareness and strengthen early detection and timely treatment to improve clinical outcomes [31].

Studies conducted in various parts of the world have identified important predictors for late-stage CC diagnosis, although there are inconsistencies among these reports. These commonly identified factors include rural residence, older age, HIV infection, high parity, low socioeconomic status, out-of-pocket expenditure, lack of cervical cancer screening practice, and low literacy status. [10, 13, 14, 32, 33]. The current study indicated that place of residence, HIV serostatus, anemia (Haemoglobin in mg/dl<12), and cervical mass size were significantly associated with CC stage at presentation. Rural dwellers had higher odds of being diagnosed with late-stage CC. More than three-quarters (76.9%) of patients with late-stage CC were from rural settings. This is in line with findings from Wassie. et.al [16] and a systematic review by Tekalign et.al [15]. This might be because patients who live in rural areas may lack information about the symptoms and signs of cervical cancer, as well as lack access to cervical cancer screening, diagnosis, and treatment services. Social, cultural, and economic barriers to CC screening services may accompany this [34].

In this study, Anaemia was found to be a strong predictor of late stage at CC diagnosis. About 41% of women with late-stage CC had a haemoglobin level less than 12 gm/dl at the time of diagnosis, and 78.3% of women in this group presented with vaginal bleeding. This finding is in line with studies from Addis Ababa. [16] and Tanzania [35], where anaemia was found to be a predictor of late-stage CC diagnosis. The link between anaemia and late-stage CC may be explained by the fact that patients with advanced-stage cervical cancer may have vaginal bleeding for a prolonged period due to tumour-related bleeding. There is also a demand for haemoglobin by the red blood cells in the formation of new blood vessels in a process called angiogenesis, with a resultant decrease in haemoglobin concentration[36].

We also found that patients with HIV were less likely to have late-stage CC at diagnosis. Studies have recognized HIV as a risk factor for late-stage CC at diagnosis [10, 37], yet other studies have reported no association [38–40]. The finding in the current study may be because of the integration of CC screening with HIV care and support; hence, these women may have been given information about the disease during their follow-up visit to antiretroviral therapy, which may improve their knowledge about cervical cancer, and therefore, may have undertaken CC screening, leading to early diagnosis. The pooled prevalence of CC screening was highest among HIV positive women in a systematic review and meta-analysis done in Ethiopia, which supports this argument.[31, 41].

This is the first study aimed at determining the stage at diagnosis of CC and identifying the factors associated with late-stage CC at diagnosis in a tertiary hospital in north-western Ethiopia. Apart from the strengths, findings should be interpreted considering the following limitations in mind. First, this was a cross-sectional population analysis in a single centre. Second, information on other key factors, including educational level, HPV infection status, and clinical parameters, was lacking. Third, incomplete information in medical records or errors in data entry may lead to misclassification. Future analyses of multicentre, prospectively collected data, including clinical and non-clinical factors, may be required to address these limitations.

## Conclusions

The current study indicated that the proportion of late-stage presentation among cervical cancer patients was high. Those patients coming from rural areas, with anaemia, larger cervical masses, and HIV positive serostatus were more likely to present with late-stage cervical cancer at diagnosis. We recommend future prospective and longitudinal studies that rigorously address questions examined in the current study. We also recommend expanding CC education for rural dwellers and cancer treatment centres, and prioritizing patients with anaemia and larger cervical masses are the areas of intervention. In addition, a qualitative approach should be incorporated to have a better understanding of the factors that may lead to a delay in seeking cervical cancer diagnosis and treatment.

## Data Availability

The minimal data set will be made available upon acceptance of the manuscript.

## Abbreviations/acronyms

BDU: Bahir Dar University
CMHS: College Medicine and Health Sciences
CC: Cervical Cancer
CT: Computed tomography
CBHI: Community-based health insurance
FIGO: International Federation of Gynaecology and Obstetrics
HIV: Human immunodeficiency virus
HPV: Human Papillomavirus
hrHPV: high-risk Human papillomavirus
IRB: Institutional Review Board
IVP: Intravenous pyelogram
MRI: Magnetic resonance imaging
OR: Odds Ratio
PET: Positron emission tomography
TGSH: Tibebe Ghion Specialized Hospital

## Acknowledgement

The research team would like also to thank Bahir Dar University College of Medicine and Health Sciences IRB who conducted ethical review and offered ethical clearance letter. We are grateful to the data collectors and supervisors involved in the study.

## Supporting information

**S1 File. Questionnaire designed to collect data**

**S1 Checklist. STROBE 2007 (v4) Statement – Checklist**

